# Design and Technical Evaluation of a Standards-Based System That Delivers Enclave-Derived Case-Finding Models to Patients and Clinicians

**DOI:** 10.64898/2026.10.20.26365982

**Authors:** Amir Asiaee, Kaveh Aryan

## Abstract

**Background:** Research data enclaves such as the All of Us Research Program do not allow participant-level data, or models fitted to them, to leave, which makes it hard to deliver *case-finding models*, which flag patients with possibly uncoded conditions for clinician review, to patients and clinicians.

**Objectives:** We describe and technically evaluate a system that fits models outside an enclave from released summary counts and delivers them through standard health information technology interfaces.

**Methods:** We exported dissemination-compliant summary counts from the All of Us Researcher Workbench and, outside the enclave, fitted versioned models for Long COVID, chronic fatigue, fibromyalgia, endometriosis, Lyme disease, lupus, migraine, depression, and anxiety. A model contract (a model file and a written specification), a Python reference implementation, and synthetic Fast Healthcare Interoperability Resources (FHIR) test patients define the expected outputs. One JavaScript scorer runs in a patient demonstration, a SMART on FHIR clinician application, and a CDS Hooks service. An atlas displays aggregate results. We evaluated the system on synthetic records with automated tests, cross-implementation output comparison, launches through the public SMART sandbox, and a counterexample review of the scoring and display rules.

**Results:** The deployed Long COVID model had an area under the receiver operating characteristic curve of 0.67 in a random validation split. The eight self-report models reached 0.61 to 0.82 in identifying self-reporters without a code. The JavaScript scorer reproduced all 162 reference outputs exactly, and all 437 automated tests passed. A counterexample review found 5 classes of failure, including scoring ineligible patients and treating unavailable data as absent, all fixed and added as tests.

**Conclusions:** Models fitted from released summary counts can be delivered through FHIR, SMART on FHIR, and CDS Hooks with reproducible scoring on synthetic records. Real-record interoperability, usability, and prospective clinical performance remain to be evaluated.

## 1. Background and Significance

Conditions such as Long COVID, myalgic encephalomyelitis/chronic fatigue syndrome (ME/CFS), fibromyalgia, and Lyme disease are often diagnosed late or not recorded with a diagnosis code. In the All of Us Research Program, the ratio of recorded Long COVID diagnoses to survey-based prevalence was about 1 in 9, assuming that All of Us participants with COVID-19 resemble the survey’s adults [1]. Among respondents reporting ongoing care for chronic fatigue with at least 5 years of electronic health record (EHR) history, the share with a chronic fatigue code was 0.28 times the mean coded share for hypertension and type 2 diabetes among respondents with the same length of EHR history [2]. *Case-finding models* flag patients whose records suggest a condition that has not been coded, so that a clinician can review them. The data needed to build these models are often stored in research enclaves.

The All of Us Researcher Workbench allows only aggregate results that meet its dissemination policy to leave the enclave, and it does not allow models trained on participant rows to be exported [3]. To meet these restrictions, a model for use outside the enclave must be constructed without exporting participant-level data or a model fitted directly to them. Published models are also often not deployed because deployment requires engineering for data access, scoring, and display inside clinical systems [4]. Standards support each step: FHIR R4 for data [5], SMART on FHIR for launching applications from the EHR [6, 7], and CDS Hooks for returning decision-support cards to the EHR [8].

## 2. Objectives

We describe the design of a system that (1) fits deployable models outside the enclave from released summary counts, (2) runs them with one scoring implementation across patient, clinician, and integration settings, and (3) uses FHIR R4, SMART on FHIR, and CDS Hooks for use with compatible EHR systems. We report a technical evaluation on synthetic records, reported following STARE-HI [9].

## 3. Methods

### 3.1. Design requirements

The design followed 5 requirements: (1) no participant-level data or model trained on those data leaves the enclave, (2) one model definition governs scoring in every application, (3) every clinician-facing output shows the chart findings behind it, (4) patient-facing screens show no model probabilities or multiples of the average risk, and (5) public demonstrations use synthetic patients only. Requirement 4 follows from what the models predict, which is a later diagnosis code or a self-reported diagnosis, not disease. A probability shown to a patient without a clinician could be read as a diagnosis or as false reassurance, so the patient sees the chart findings and the clinician sees the estimate. Figure 1 shows the architecture and data boundaries.

**Figure 1:**
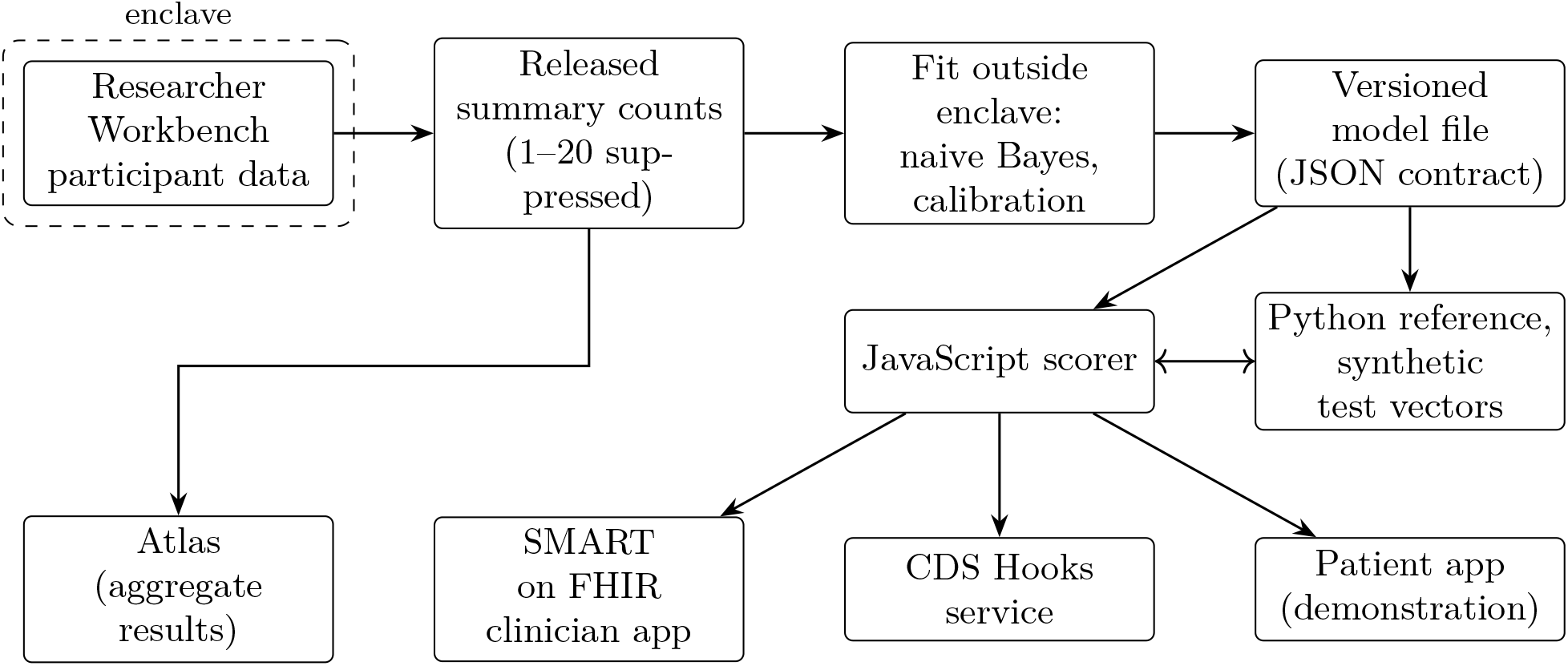
System architecture and data boundaries. Only summary counts that meet the dissemination requirements leave the All of Us Researcher Workbench (dashed boundary). We fit models outside the enclave and store them as versioned JSON files. A Python reference implementation and synthetic FHIR test patients define the expected outputs. The JavaScript scorer must reproduce these outputs (double arrow). The scorer runs in the clinician application, the CDS Hooks service, and the patient demonstration. The atlas shows aggregate results only.

### 3.2. Models fitted from released counts

The underlying analyses used the All of Us Research Program Registered Tier Dataset v9 (curated data repository R2025Q4R6, data cutoff January 1, 2025) [10]. Inside the Workbench, we suppressed counts of 1 to 20, rounded other counts up to a multiple of 10, and, within each table, suppressed a further count wherever a suppressed count could otherwise be recovered from the table’s totals (*complementary suppression*). For each candidate feature, the Workbench released a *feature-by-label table*: counts of patients with and without the feature among those with and without the label. It also released the label totals. We retained features only when every cell of their feature-by-label table exceeded 20 [3].

Outside the enclave, we fitted each model as naive Bayes. Each feature’s weight is its log odds ratio from the feature-by-label table, shrunk toward zero by empirical Bayes. Features with few patients receive more shrinkage. We then scored the training split inside the Workbench with the fitted weights and calibrated each model from the released numbers of patients with and without the label within bins of that score [1, 11]. Each analysis divided its cohort at random into a training split, from which the counts were released, and a 20% validation split.

The nine models differ in what they predict, a later diagnosis code for Long COVID and a self-reported diagnosis for the other eight. The Long COVID model predicts subsequent documentation. We define *day 0* as the first recorded COVID-19 diagnosis (ICD-10-CM U07.1) on or after July 2, 2021, as in the training cohort. Predictors use records through day 90, the outcome is a first U09.9 code in days 91 to 455, and the application can score a patient on any *scoring date* from day 90 to day 455. A *qualifying code* is the target condition’s own diagnosis code, listed in each model file. The other eight models (chronic fatigue, fibromyalgia, endometriosis, Lyme disease, lupus, migraine, depression, and anxiety) predict a self-reported prior diagnosis among adults with no qualifying code (for example, systemic lupus erythematosus [M32] for lupus and A69.2 for Lyme disease), from records in the 5 years before the scoring date. The endometriosis model covers women only. The Lyme disease and lupus models add condition-specific findings, tests, and treatments to the same feature set. The chronic fatigue label does not establish ME/CFS. The Lyme disease label mostly reflects past, treated infection. We have not validated any self-report model against an independent assessment of current disease. The *development-cohort label prevalence* is the proportion of participants with the label in that cohort, which the application reports as the average risk. *Risk bands* (Typical, Elevated, High) are heuristic display thresholds at 1.5 and 3 times this average.

### 3.3. Model contract and test vectors

The *model contract* consists of the JSON model file and a written specification of how to evaluate it on a FHIR R4 bundle. The file contains the features with their codes and windows, weights, intercept, calibration map, risk bands, and eligibility rules. The written specification defines which resources and codes count for each feature, where windows fall relative to day 0 or the scoring date, when not to score a patient, and how the score maps to a probability, a band, and a multiple of the average. The scorers evaluate only patients whom the training cohort would have included. They count only records dated on or before the scoring date. When a FHIR search fails, they return an explicit could-not-score result, rather than treating unavailable records as absent findings. Supplementary Section S5 lists these rules.

The Python reference implementation defines the expected software outputs. Eighteen synthetic FHIR patients, each built to exercise specific rules (10 demonstration patients and 8 eligibility boundary cases from the counterexample review in Section 3.6), provide the test cases. A *test vector* pairs one synthetic patient’s FHIR bundle with the reference output for one model. The JavaScript scorer is a function-by-function translation of the Python reference implementation. It must reproduce the reference output for every test vector, including the order of findings. Each scored result lists the contributing features and the supporting chart records. Figure 2 applies one model file to one synthetic chart.

**Figure 2:**
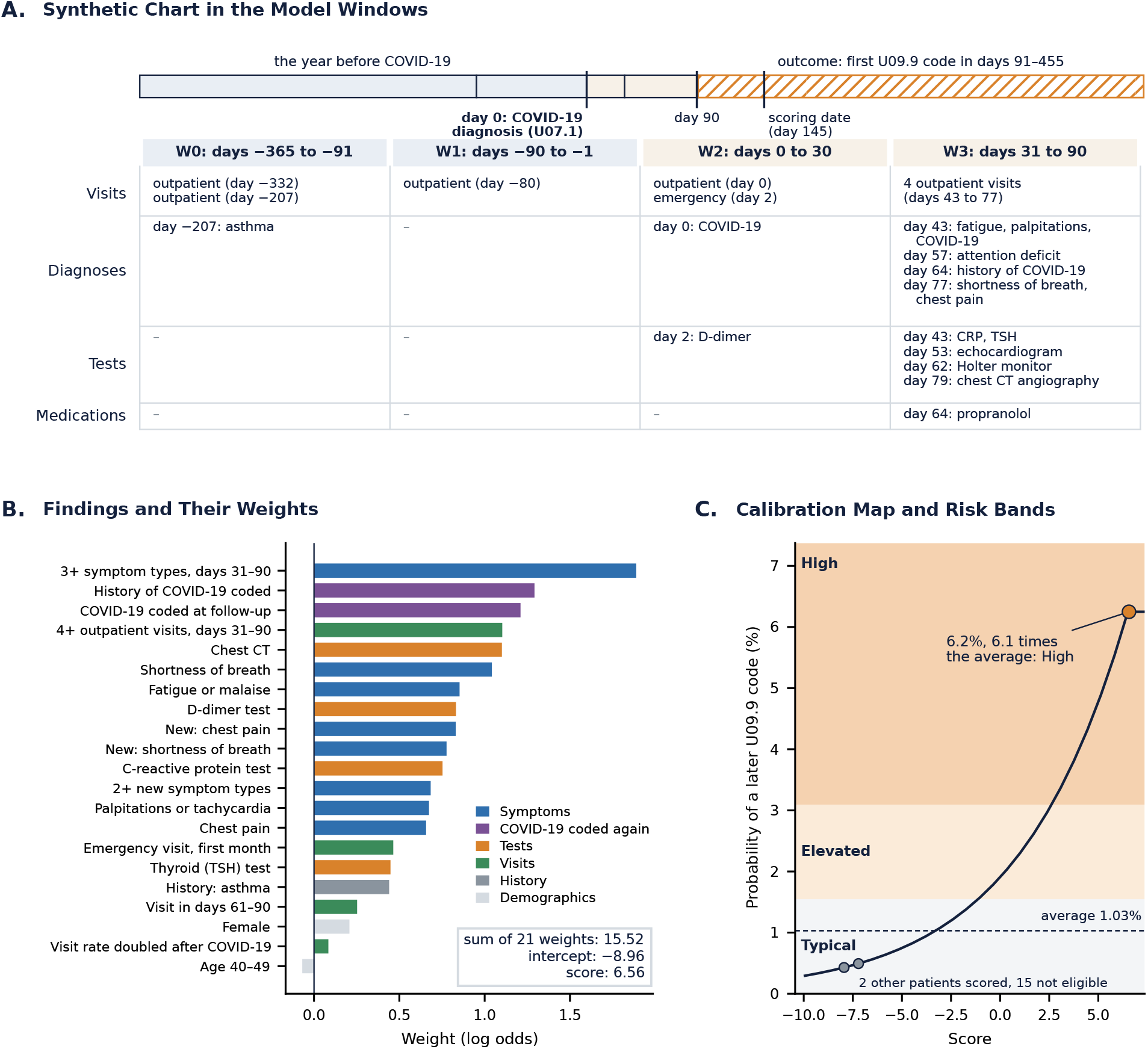
One model file applied to one synthetic chart. **A.** The chart of a synthetic 43-year-old woman, arranged by the Long COVID model’s windows relative to her COVID-19 diagnosis (day 0). Only records dated on or before the scoring date count, and the model predicts a first U09.9 code in days 91 to 455. **B**. The 21 model features present in the chart, each with its weight. A feature’s weight is its contribution to the score in log odds before calibration. The score is the intercept plus the sum of the weights. **C**. Because naive Bayes treats correlated findings as independent, its score overstates certainty, and the calibration map, fitted to the training split’s observed labels, converts the score to probability. The risk bands are at 1.5 and 3 times the 1.03% average. Scores beyond the last point of the map receive the probability at that point. Gray points are the other 2 synthetic patients the model scores. CRP, C-reactive protein. TSH, thyrotropin.

### 3.4. Components

#### Patient demonstration

Several of these conditions are diagnosed from symptoms that charts often miss, such as post-exertional malaise or widespread pain. The patient demonstration lets patients report those symptoms so that a clinician sees them beside the chart findings, and for Long COVID, ME/CFS, and fibromyalgia, a questionnaire result also raises or lowers the chart estimate. Patients answer one questionnaire in an installable web application (Figure 3A). It combines published instruments: the 2024 RECOVER-Adult symptom index for Long COVID [12], with item wording from the RECOVER questionnaire [13], the DePaul Symptom Questionnaire post-exertional malaise items for the defining symptom of ME/CFS, which also occurs in Long COVID [14], and the Post-COVID-19 Functional Status scale for how much COVID-19 limits daily life [15].

**Figure 3:**
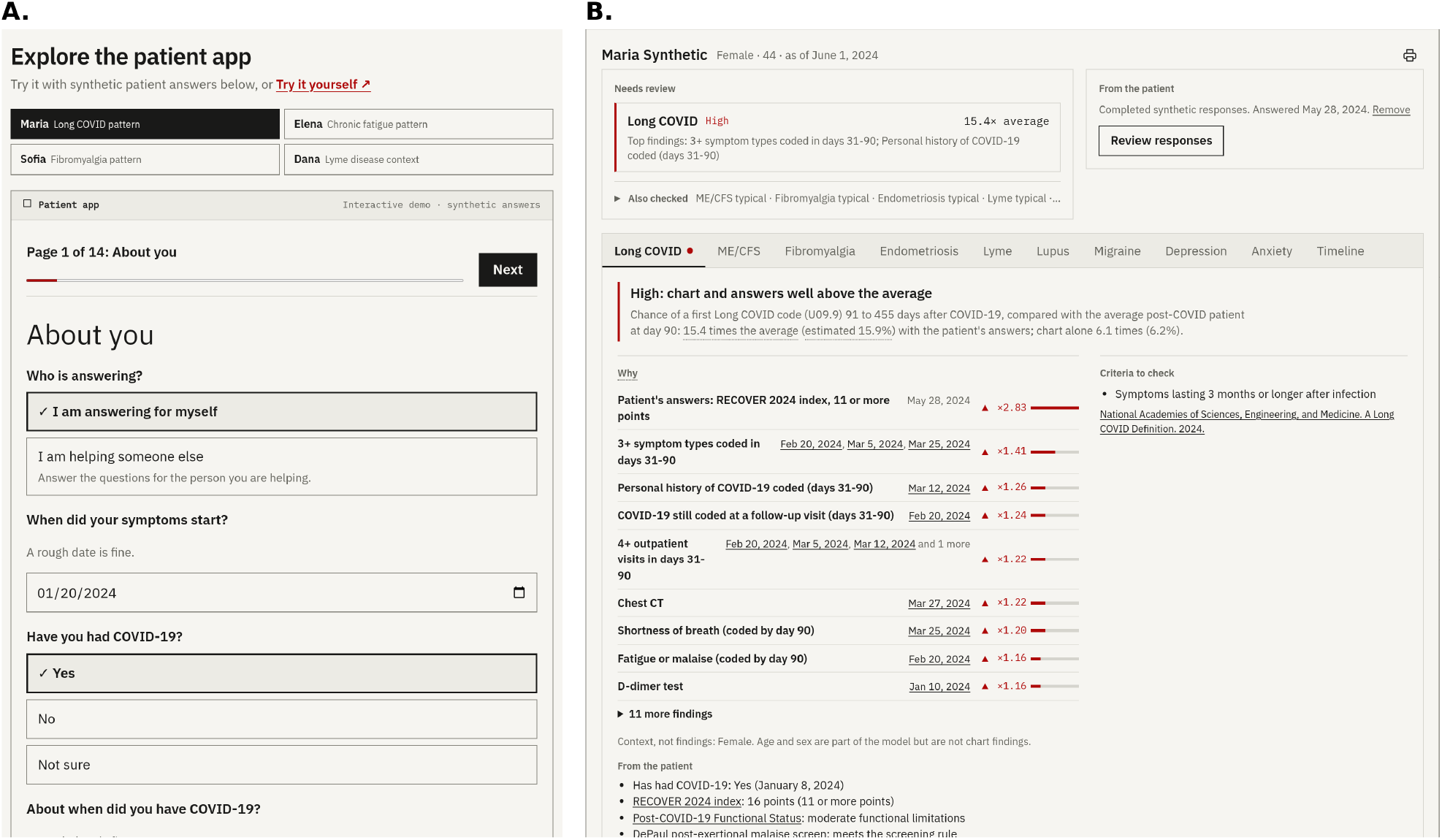
Patient and clinician views for synthetic patients. **A.** Patient demonstration with its demonstration patients and the first page of the symptom check, showing one patient’s demonstration answers. **B**. Clinician dashboard for a synthetic patient with a High Long COVID pattern, with the patient’s demonstration answers attached. The dashboard lists conditions needing review and the patient’s answers. The Long COVID tab shows the questionnaire result and each dated finding, with how much each changes the calibrated estimate.

Optional parts add condition self-checks written from published criteria for Long COVID symptom duration, ME/CFS, fibromyalgia, and possible Lyme disease exposure [16–18], the Patient Health Questionnaire-9 (PHQ-9) for depression [19], and the Generalized Anxiety Disorder-7 (GAD-7) for anxiety [20]. The application shows chart findings from the demonstration patient’s synthetic FHIR records in plain language without probabilities. Questionnaire answers are a separate input, stored in the browser. Patients can export the answers as a FHIR R4 QuestionnaireResponse that the clinician application can import. The application works in English and Spanish and offline after loading. Any answer other than “not at all” to the PHQ-9 item on thoughts of death or self-harm shows the 988 Suicide and Crisis Lifeline at once. Supplementary Section S1 gives the scoring rules, translations, and visit summary.

#### Clinician application

The clinician application brings the models into the EHR, where a clinician reviews a chart, and shows which conditions may be present but uncoded and the evidence behind each. A SMART on FHIR application, launched from the EHR with the authorization code flow and Proof Key for Code Exchange, reads the Condition, Encounter, Observation, Medication-Request, and Procedure resources that the models need and scores every model. Its dashboard, the *condition review*, lists the conditions in the High or Elevated band above a summary of the conditions checked (Figure 3B). Each condition’s tab shows the band, the dated findings behind it, the published criteria to review [16–18], and next steps. For each finding, the dashboard compares the calibrated estimates with and without that finding’s weight and displays the comparison as an odds ratio. When the imported answers yield a positive or negative result for an instrument with a published likelihood ratio measured against a reference standard similar to what the model predicts, the dashboard multiplies the odds of the chart estimate by the ratio for that result, assuming that the result is independent of the chart findings given the condition, and shows the chart-only estimate beside the combined one [21–23]. For the RECOVER index, the reference standard is a U09.9 code. The Lyme disease and lupus tabs show, beside the model’s estimate, the evidence of a chart rule built on surveillance data and testing guidance [24, 25] or on the 2019 lupus classification criteria [26]. The lupus rule also reads the text of clinical notes and reports and sums the criteria’s weights as points. When required records cannot be read, the card says “Could not score” and names the unavailable records. A no-login demonstration runs the same code on 4 of the 10 demonstration patients. Supplementary Section S2 gives the import rules, likelihood ratios, and chart rules.

#### Integration service

The integration service delivers the same results inside the EHR’s own screens, without a separate application. The EHR calls the CDS Hooks *patient-view* hook when a clinician opens a patient’s chart. The service declares *prefetch templates*, FHIR queries that the EHR runs to retrieve the resources the models read. The EHR sends these resources with the request. The service validates each request, including that every supplied or fetched resource belongs to the requested patient. It fetches missing resources with the access token included by the EHR, only from FHIR servers on a configured allow-list (by default, the public synthetic sandbox). It returns a card for a condition in the Elevated or High band. When records a model needs cannot be read, it returns one information card naming the conditions it could not score instead of an empty card list. This synthetic-data prototype does not authenticate callers. Authentication is required for deployment with real patient data. CDS Hooks 2.0 specifies authentication through a signed JSON Web Token from the CDS client [8].

#### Atlas

The atlas serves people who plan care rather than treat one patient, such as health system leaders and public health staff, by showing for whom and where each condition goes uncoded. A public page shows aggregate results for one condition at a time. It shows how often the condition is coded by age group, sex, and race and ethnicity. It also shows state maps where they were released, the coded findings more common in people with the condition, and, for chronic fatigue, fibromyalgia, and endometriosis, Fitbit activity and sleep. It draws on the aggregate releases of the analyses that developed the models [1, 2, 11] and on public survey and surveillance data [24, 27]. It shows shares and intervals, never All of Us counts. The build of its data checks that no suppressed group can be recovered from the groups shown. It does not run the scorer. Its illustrative *scenario calculator* uses assumptions entered by the user, such as the share of patients flagged and the share with a positive questionnaire result, to show how many patients a clinic would review and find. Supplementary Section S3 describes its panels and the calculator.

#### Developer documentation

The developer documentation lets another team connect an EHR to the system, or check the scorer, without our help. A documentation page describes each integration path with request and response examples produced by the running code. It runs the scorer in the browser on any test patient and compares the result with the reference output. The page also lists the model files with their hashes.

### 3.5. Data flow and privacy

Patient answers are stored locally in the browser, and the clinician application processes retrieved records in the browser. The CDS service processes supplied chart data for each request and does not store or log those data at the application level. Retention and logging by the hosting infrastructure require separate verification. Each page has a content security policy that restricts which origins it may contact. Clinician-facing outputs show supporting information for independent review. We did not determine the regulatory status of the software’s individual functions.

### 3.6. Technical evaluation

We evaluated the system on synthetic records only. We ran the automated test suites of both scorers and compared every reference output between the two implementations. We also tested synthetic CDS Hooks requests and performed structural FHIR conformance checks of the test bundles, the example requests, and the exported QuestionnaireResponses. We launched the clinician application through the public SMART Health IT sandbox and compared its condition reviews with those that the demonstration’s code produces for the same charts, for all 18 synthetic patients. Because the live application scores as of the current date, we shifted the dates of the sandbox charts so that the scoring date is the upload date.

Before these checks, we ran a *counterexample review*. It asked whether the software followed the specification and the training cohort’s definitions at the *boundary with real records*, where charts are incomplete, partial, or unusual in ways that the developers’ test patients might not cover. A reviewer working apart from development read the specification and the code and constructed synthetic inputs on which the software’s output would contradict the specification or the cohort definitions. Examples included a child given the chart of an adult who scores High, a birth date recorded only as a year, a FHIR search that returns an error, a clinical note that says “no oral ulcers,” and calculator inputs that imply more patients found than reviewed. We fixed each failure the counterexamples exposed and added every counterexample to the test suite.

## 4. Results

### 4.1. Models

Table 1 lists the *deployed models*, meaning the model files the system runs. The deployed Long COVID model contains 69 features, of which 57 have nonzero weights. It had an area under the receiver operating characteristic curve (AUROC) of 0.674 (95% CI, 0.592–0.747) in a random validation split. We did not externally validate the deployed model or its probability map. The closest evidence comes from the analysis that developed the model, which also released a linear discriminant on 41 features from the same *feature registry*, the list of features with their codes and windows. Fitted without data from 18 held-out sites, defined by the EHR source of each patient’s COVID-19 diagnosis, the linear discriminant had an AUROC of 0.643 (95% CI, 0.553–0.719) at those sites [1].

**Table 1:** Models the system runs. We fitted all models as naive Bayes with empirical-Bayes shrinkage outside the enclave from released summary counts and calibrated them from released score bins. Validation is retrospective, in the All of Us Research Program Registered Tier Dataset v9. For the self-report models, the validation AUROC compares self-reporters with no qualifying code observed after the survey (positives) with all other participants in the validation split (negatives). AUPRC, area under the precision-recall curve.

| Model (version) | Label and target | Features | Validation: AUROC (95% CI), AUPRC |
| --- | --- | --- | --- |
| Long COVID (1.2.0) | First U09.9 code in days 91–455 among adults uncoded at day 90 | 69 (57 nonzero) | Random 20% split: 0.674 (0.592–0.747), 0.028. Released linear discriminant (41 features) at 18 held-out sites: 0.643 (0.553–0.719), 0.021 |
| Chronic fatigue (1.0.0) | Self-reported diagnosis among adults with no qualifying code | 73 | Random 20% split: 0.735 (0.720–0.749), 0.156 |
| Fibromyalgia (1.0.0) | Self-reported diagnosis among adults with no qualifying code | 80 | Random 20% split: 0.726 (0.710–0.743), 0.080 |
| Endometriosis (1.0.0) | Self-reported diagnosis among women with no qualifying code | 90 | Random 20% split: 0.610 (0.595–0.621), 0.111 |
| Lyme disease (1.0.0) | Self-reported diagnosis among adults with no qualifying code | 86 | Random 20% split: 0.651 (0.632–0.673), 0.045 |
| Lupus (1.0.0) | Self-reported diagnosis among adults with no qualifying code | 79 | Random 20% split: 0.824 (0.777–0.863), 0.029 |
| Migraine (1.0.0) | Self-reported diagnosis among adults with no qualifying code | 88 | Random 20% split: 0.691 (0.680–0.700), 0.305 |
| Depression (1.0.0) | Self-reported diagnosis among adults with no qualifying code | 86 | Random 20% split: 0.759 (0.751–0.766), 0.477 |
| Anxiety (1.0.0) | Self-reported diagnosis among adults with no qualifying code | 84 | Random 20% split: 0.792 (0.782–0.802), 0.442 |

The chronic fatigue and fibromyalgia models had AUROCs of 0.735 and 0.726 in random validation splits. The Lyme disease and lupus models had AUROCs of 0.651 and 0.824. The endometriosis model, scored only for women, reached 0.610. Its discrimination is limited, and the ever-diagnosed survey label may not represent current disease. Its clinician card states these limitations. Models for migraine, depression, and anxiety, fitted the same way, reached 0.691, 0.759, and 0.792. They are published as model files with reference outputs. Like the other models, they are shown in the clinician application and returned by the CDS Hooks service.

### 4.2. One synthetic patient through the system

Figure 2 follows one synthetic patient, a 43-year-old woman with asthma, through the Long COVID model. Her COVID-19 diagnosis on January 8, 2024, is day 0. In the next 90 days her chart records an emergency visit with a D-dimer test and 4 outpatient visits. It also records fatigue, palpitations, attention deficit, shortness of breath, and chest pain. Tests and treatments include C-reactive protein and thyroid tests, an echocardiogram, a Holter monitor, chest computed tomography angiography, and propranolol (Figure 2A). Twenty-one model features are present. Their weights sum to 15.52 log odds. Adding the intercept gives a score of 6.56 (Figure 2B). Her score lies just beyond the last point of the calibration map, so the map gives her its highest probability, 6.2%, of a first U09.9 code in days 91 to 455. This probability is 6.1 times the 1.03% average and falls in the High band (Figure 2C). Of the other 17 synthetic patients, 2 score Typical and 15 are not scored. Each unscored result gives a reason, such as a Long COVID code already recorded or no qualifying COVID-19 diagnosis. The clinician application lists Long COVID among the conditions needing review, with the same dated findings (Figure 3B). The CDS Hooks service returns a warning card with the same estimate. The patient demonstration shows her chart findings in plain language without the probability.

### 4.3. Technical evaluation

Table 2 summarizes the outcomes. All 437 JavaScript tests and the Python reference tests passed. The two scorers agreed on all 162 reference outputs (18 synthetic patients under each of the 9 models). The synthetic CDS Hooks requests returned the expected cards. The structural conformance checks passed. In the public SMART sandbox, the clinician application gave the same condition review as the demonstration for all 18 patients after we corrected 5 defects that the comparison exposed, 3 in the tool that loads the synthetic charts into the sandbox and 2 in the application (Section 5). Separately, the counterexample review found 5 classes of failure (Table 3). Three of them, eligibility, partial dates, and unavailable data, concern how the scorer reads a real chart. The 10 demonstration patients did not cover these cases.

**Table 2:** Technical evaluation results on synthetic records. CDS, clinical decision support. HL7, Health Level Seven.

| Check | Outcome |
| --- | --- |
| Automated tests (Node.js, Python) | All 437 JavaScript tests and the Python reference tests passed |
| Cross-implementation parity | 162 of 162 reference outputs reproduced (18 patients, 9 models) |
| CDS Hooks (synthetic requests) | Discovery and patient-view requests returned the expected cards |
| SMART launch (public sandbox) | All 18 condition reviews matched the demonstration after 5 defects were fixed |
| Counterexample review | 5 failure classes found (eligibility, partial dates, unavailable data, lupus rule extraction, and calculator feasibility, Table 3), all fixed and added as tests |
| FHIR conformance | Structural checks passed for all bundles, example requests, and QuestionnaireResponses |

**Table 3:** Failure classes found by the counterexample review. Each row shows a counterexample for a failure class and the behavior before and after the fix. Every counterexample is included in the synthetic test suite. CDS, clinical decision support.

| Failure class | Counterexample and the failure it exposed | Behavior of the system |
| --- | --- | --- |
| Eligibility | A child given the chart of an adult who scores High was scored. A Long COVID code dated before the COVID-19 diagnosis did not exclude the patient | Patients younger than 18 years or with a code for the condition by the scoring date, and for Long COVID patients outside days 90 to 455, are not scored, and the card says why |
| Partial dates | A birth date recorded only as a year (1980) stopped scoring, and the CDS Hooks service returned no cards | A partial record date reads as the earliest compatible day, and a partial birth date as July 1 or the 15th of the month, as in the training cohort |
| Unavailable data | A forbidden Observation search became an empty result, so its findings counted as absent. The CDS Hooks service could return an empty card list | A failed, forbidden, or truncated search gives “Could not score,” and the CDS Hooks service returns an information card |
| Lupus rule extraction | “Subacute cutaneous lupus” also matched “acute cutaneous lupus” (6 points instead of 4). “No oral ulcers. No seizure. No pericarditis.” gave 13 potential points. White-cell counts of $3 \times 10^9/\text{L}$ and $3,000/\mu\text{L}$ gave different results | Longest matches on word boundaries, negated mentions ignored within the clause, and counts compared in $10^9/\text{L}$ |
| Calculator feasibility | A 1% share of patients with a positive questionnaire result gave 5 patients reviewed and 16 found, a precision of 320% | A dependent input is raised to its nearest feasible value, and the page says so |

## 5. Discussion

This prototype shows that models fitted outside a research enclave from released summary counts can be packaged as a versioned contract and delivered through FHIR, SMART on FHIR, and CDS Hooks. The JavaScript scorer reproduced the reference outputs exactly on synthetic records. We contribute the route from an enclave to clinical interfaces. The analyses that developed the models report their discrimination in detail [1, 11].

Separating the model contract from the applications simplified testing and model integration. The test vectors let us check the JavaScript scorer against the reference without real patient data.

Adding a model requires no change to application code, only a model file and its test vectors. Adding a new condition requires a feature registry, a compliant aggregate extraction, model fitting and calibration, implementation checks, and condition-specific validation. Compared with deployment frameworks that run custom models inside one health system’s infrastructure [4], the contract here is static and portable. This design suits enclave-derived models but restricts them to binary features with sufficient counts.

The enclave’s output rules constrained the choice of model. Because no model fitted to participant rows may leave the enclave, shared models must be fitted outside it from the released feature-by-label counts and score bins alone. These summaries support fitting naive Bayes and linear discriminants [1], but not models that require participant rows, such as gradient-boosted trees. Running a richer model inside the enclave and returning its scores would not help a clinic, because the patients a clinician sees are not in the enclave. Naive Bayes also serves design requirement 3. Each weight is a per-feature log odds ratio, so the application can show how much each dated finding changes the estimate (Figure 2B). In exchange, the models have modest discrimination and use only binary features.

Unreadable data require an explicit result. A FHIR search can fail, be forbidden, or return only part of a record. A scorer that treats unavailable records as absent findings can give a falsely reassuring estimate (Table 3). CDS Hooks makes partial failures hard to report. A service that cannot retrieve the FHIR data it needs can respond with HTTP status 412 (Precondition Failed), which withholds every result [8]. A service that scores some models but not others has no such signal. The card format has no field for a partial result, and a condition without a card looks the same whether its model found nothing to flag or could not score the patient. Our service therefore returns one information card that names the conditions it could not score. A standard way to report a partial evaluation would help any CDS Hooks service whose output depends on complete records.

Real FHIR servers also behave differently from test files. The public SMART sandbox splits search results into pages and returns them in its own order. Our application, tested on single bundles, did not follow the sandbox’s next-page links. The server’s ordering also determined which dated record a finding cited. Scoring also depends on the dates in the synthetic charts. The application scores as of the current date, so a synthetic chart keeps the demonstration’s result only while its records stay inside the model’s windows, and the charts must be re-dated each time they are loaded. Teams that build time-windowed models into SMART on FHIR applications should test them against a server that splits results into pages and reorders them. The synthetic data should be dated relative to the test date.

The counterexample review showed that passing our own tests did not guarantee correct behavior at the boundary with real records. Eligibility rules, partial dates, and unavailable data are where a deployed tool meets real records. Each counterexample is included in the test suite to detect a recurrence of that failure. The suite covers only the cases someone thought to construct. A reviewer who did not write the code used the specification to construct inputs on which the software might contradict it. This review found failures that our own tests had missed. We recommend asking a reviewer to construct such counterexamples for similar systems.

Other enclaves could reuse the route if they release feature-by-label counts and score bins under small-cell rules. The model contract, the reference implementation, and the scorer would carry over unchanged only if the other enclave’s feature registry uses the same standard codes (ICD-10-CM, CPT, LOINC, RxNorm). The receiving EHR must also expose FHIR R4 with SMART on FHIR or CDS Hooks. The next step is to run the scorers on real records at a site with FHIR access and compare the features they compute with those computed from the research data model. This comparison should precede any evaluation with clinicians or patients.

### 5.1. Limitations

This evaluation has four main limitations. First, it used synthetic records only, so we did not evaluate real-record interoperability, usability, or clinical performance, and we did not test launches from commercial EHRs. Second, the models predict a later diagnosis code or a self-reported diagnosis rather than independently established disease. Their scores may reflect care access, utilization, and coding practices. We validated them retrospectively in one cohort and have not evaluated their calibration, the heuristic risk bands, or the questionnaire likelihood ratios in deployment populations. Third, we have not shown that features computed from FHIR records equal those computed from the research data model. Participant-provided FHIR data in All of Us hold far fewer condition and procedure records than the research data [28]. The application reads FHIR administrative gender rather than sex at birth. Fourth, the CDS service does not authenticate callers, which a deployment with real patient data requires. Supplementary Section S4 lists narrower limitations of individual components.

## 6. Conclusion

This prototype combines models fitted from released summary counts with a shared scoring implementation, patient and clinician interfaces, a CDS Hooks service, and an aggregate-data atlas. Synthetic testing established reproducible scoring across the reference and JavaScript implementations. It also identified 5 failure classes at the boundary with real records. We corrected these failures and included tests for each class. Real-record interoperability, usability, and prospective clinical performance remain to be evaluated.

## Supporting information

STARE-HI Checklist

## Data Availability

All of Us Registered Tier Dataset v9 is available to authorized researchers through the All of Us Researcher Workbench. The source code, model files, test vectors, released aggregate counts, and the scripts that refit the models and rerun the automated checks are available at https://github.com/AsiaeeLab/unom-system. A live demonstration with synthetic patients is at https://undiagnosednomore.org.

## Clinical Relevance Statement

This prototype delivers models fitted from released summary counts to patient and clinician interfaces. Testing established technical consistency on synthetic records, while clinical effectiveness remains untested. Real-record interoperability, usability, and prospective clinical evaluation are needed before routine use.

## Multiple-Choice Questions

1. Why were the models fitted outside the enclave? (a) Improved accuracy from training on summary counts. (b) Compliance with enclave export restrictions. (c) Compliance with FHIR requirements for naive Bayes. (d) Automatic calibration from training on summary counts. *Correct answer: (b)*. The All of Us Researcher Workbench allows only aggregate results that meet its dissemination policy to leave. A model fitted outside from released counts can be shared.
2. What did the cross-implementation test vectors establish? (a) Clinical validity of the models. (b)Agreement of the scorer with the reference outputs on synthetic patients. (c) Interoperability with real patient records in commercial EHRs. (d) Calibration of the risk bands in deployment populations. *Correct answer: (b)*. The vectors check software consistency. The study did not evaluate clinical validity, EHR interoperability, or calibration in deployment.

## Protection of Human and Animal Subjects

The underlying analyses used de-identified data from the All of Us Research Program Registered Tier Dataset v9 (curated data repository R2025Q4R6, data cutoff January 1, 2025). All of Us participants provide informed consent at enrollment under a protocol overseen by the program’s institutional review board. We analyzed de-identified Registered Tier data as authorized researchers [10]. The system evaluation used synthetic patients only and involved no human subjects.

## Acknowledgments

We gratefully acknowledge All of Us participants for their contributions, without whom this research would not have been possible. We also thank the National Institutes of Health’s All of Us Research Program for making available the participant data examined in this study. The All of Us Research Program is supported by the National Institutes of Health, Office of the Director: Regional Medical Centers: 1 OT2 OD026549; 1 OT2 OD026554; 1 OT2 OD026557; 1 OT2 OD026556; 1 OT2 OD026550; 1 OT2 OD 026552; 1 OT2 OD026553; 1 OT2 OD026548; 1 OT2 OD026551; 1 OT2 OD026555; IAA #: AOD 16037; Federally Qualified Health Centers: HHSN 263201600085U; Data and Research Center: 5 U2C OD023196; Biobank: 1 U24 OD023121; The Participant Center: U24 OD023176; Participant Technology Systems Center: 1 U24 OD023163; Communications and Engagement: 3 OT2 OD023205; 3 OT2 OD023206; and Community Partners: 1 OT2 OD025277; 3 OT2 OD025315; 1 OT2 OD025337; 1 OT2 OD025276. In addition, the All of Us Research Program would not be possible without the partnership of its participants.

## Data and Code Availability

This study used data from the All of Us Research Program’s Registered Tier Dataset v9, available to authorized users on the Researcher Workbench. A public repository (MIT License), at https://github.com/AsiaeeLab/unom-system, holds the source code, the model files, the test vectors, the released counts, and the scripts. These scripts refit every model file, rebuild Table 1, and rerun the automated checks of Table 2. A live demonstration with synthetic patients runs the same code at https://undiagnosednomore.org.

