## Supplementary Material for "Design and Technical Evaluation of a Standards-Based System That Delivers Enclave-Derived Case-Finding Models to Patients and Clinicians"

This supplement gives the component details that the main text summarizes in Section 3.4 and the scoring rules that Section 3.3 summarizes, and the limitations of individual components that Section 5.1 refers to. Figure and table numbers refer to the manuscript.

### S1 Patient demonstration

*Instruments.* The installable web application administers the 2024 RECOVER-Adult symptom list [1], with item wording from the RECOVER questionnaire [2], the DePaul Symptom Questionnaire post-exertional malaise items [3], the Post-COVID-19 Functional Status scale [4] (for people who report possible or prior COVID-19), and 5 custom change questions. The app shows the RECOVER total and its research category only when every symptom is answered and otherwise shows the points so far as incomplete.

*Optional parts.* Condition self-checks, written for the app from published criteria, cover Long COVID symptom duration (the 2024 National Academies definition [5]), the ME/CFS core symptoms [6], the fibromyalgia Widespread Pain Index and Symptom Severity Scale [7], and possible Lyme disease exposure. Each reports whether the answers match the pattern the criteria describe and is a prompt for the clinician, not a diagnosis. Two further parts give the published Patient Health Questionnaire-9 (PHQ-9) [8] and Generalized Anxiety Disorder-7 (GAD-7) [9] totals and severity bands once every item is answered. Any answer other than “not at all” to the PHQ-9 item on thoughts of death or self-harm shows the 988 Suicide and Crisis Lifeline at once, also offline, and puts a notice at the top of the patient’s answers in the clinician view.

*Languages.* The app runs in English or Spanish, storing the same answers and computing the same scores in either language. The Spanish version uses the RECOVER study’s Spanish survey wording, the scale authors’ Spanish translation of the functional status scale, and Spanish versions of the DePaul items published by the questionnaire’s developers, and we translated the remaining questions for the app.

*Records, storage, and sharing.* Answers are stored only in the browser. The demonstration shows findings from synthetic records in plain language, with dates and without model probabilities or risk multiples. Connection to a person’s own health record remains planned. A printable visit summary lets patients choose what to share, and questionnaire answers can be downloaded as a FHIR R4 QuestionnaireResponse with stable item identifiers or handed to the clinician demonstration within the browser. After initial loading and caching, the questionnaire works offline, but trial searches use the ClinicalTrials.gov API and need a network connection.

#### S2 Clinician application

*Dashboard and condition tabs.* The SMART on FHIR application reads the patient's conditions, encounters, observations, medication requests, and procedures. The dashboard lists conditions needing review (High or Elevated bands) above a one-line summary of the conditions checked and shows the patient's questionnaire answers and self-check results when present. Each condition has a tab with the band and up to 8 dated findings (more on expansion). Each finding is shown with the odds ratio between the estimate with and without it, on one scale shared by every patient and model (a full bar is a doubling or halving of the odds), and findings that lower the estimate are included. Demographic inputs are listed separately. The tab also gives the criteria to review (the 2024 National Academies Long COVID definition [5], the Institute of Medicine 2015 ME/CFS criteria [6], and the 2016 revised fibromyalgia criteria [7]), next steps, and a timeline of chart events. A card whose model could not read the records it needs says "Could not score" and names the missing records, and the summary then notes that a result is missing.

*Questionnaire import and combination.* The dashboard imports only responses to the app's own Questionnaire in a supported version and for the open patient, selects the most recent valid one, and attaches a response that names no patient only after the clinician confirms it. When the attached response has a result for a condition's instrument, the dashboard updates the chart estimate by Bayes' rule, multiplying the chart odds by that result's published likelihood ratio, measured against a reference close to the model's target: the RECOVER index against a U09.9 code (2.83 if positive, 0.46 if negative) [10], questionnaire-based Institute of Medicine criteria against a physician panel's diagnosis among fatigued adults (1.62, 0.47) [11], and the patient-completed 2016 fibromyalgia criteria against a reported physician diagnosis (8.46, 0.70) [12]. The band, multiple of the average, and next steps then follow the combined estimate, with the chart-only values shown beside it. Instruments without a published likelihood ratio against a comparable reference (the post-exertional malaise screen alone, the Long COVID time criterion, the functional status scale, the Lyme exposure questions, the PHQ-9, and the GAD-7) are shown but not combined.

*Chart rules.* The Lyme disease and lupus tabs show their model's estimate beside the evidence of a chart rule. For Lyme disease, the rule lists coded findings consistent with the infection, with historical surveillance incidence for the patient's county [13] and CDC testing guidance [14]. For lupus, it maps chart evidence to the 2019 classification criteria [15], matching criterion terms on word boundaries, counting only the longest of overlapping matches, ignoring negated mentions in the same clause, and comparing white-cell and platelet counts in a common unit. The no-login demonstration runs the same code on 4 synthetic personas drawn from the 10 demonstration patients of the test corpus.

#### S3 Atlas

The atlas shows one condition at a time (Long COVID, ME/CFS, Lyme disease, lupus, fibromyalgia, migraine, endometriosis, depression, or anxiety): a state map where one was released, how often the condition is coded by age group, sex, and race and ethnicity, and the coded findings more common in people with the condition. For the self-reported conditions, coding is the share of people who report the diagnosis and have its code, shown beside hypertension and type 2 diabetes in the same group. For Long COVID, coding is the coded share relative to survey prevalence, and the map adds state prevalence from the Household Pulse Survey [16]. The Lyme disease map adds county incidence from CDC surveillance [13]. For ME/CFS, fibromyalgia, and endometriosis, a panel compares Fitbit steps, sedentary and very active minutes, and sleep in the year before the survey between people who report the condition and people who do not, standardized to the same age and sex mix.

For Long COVID, a scenario calculator uses comparator-model inputs and user-specified assumptions. When the inputs would imply an impossible yield, such as more patients found than reviewed, it raises the dependent input to the nearest feasible value and says so. The calculator's outputs are illustrative.

#### S4 Limitations of individual components

*Combining the questionnaire with the chart.* The update with the patient's answers treats them as independent of the chart findings given the condition, as the naive Bayes models treat their own features. Where the chart codes the same symptoms the answers count, the combined estimate overstates. None of the likelihood ratios was measured in the application's population, and because the app's RECOVER total can only exceed the research index, the published Long COVID ratio likely overstates the app's.

*Questionnaire.* The RECOVER items use the published questionnaire's symptom wording [2], but the question stem is ours. The app does not administer the severity measures that the index requires for fatigue, brain fog, and chest pain, so its total can exceed the research index by up to 5 points. The combined questionnaire and chart triage rule and the condition self-checks are exploratory. We restricted the Post-COVID-19 Functional Status scale to participants who report possible or prior COVID-19, and the custom change questions use symptom onset when COVID-19 does not apply. We have not validated the resulting questionnaire workflow for chronic fatigue case finding.

*Conformance and security.* Our own tests checked FHIR conformance structurally, and we did not run the HL7 FHIR validator. Records hidden by security labels are treated as readable rather than unavailable.

*Lupus rule.* The rule detects negation with a short list of cues, and no rheumatologist has reviewed its outputs.

#### S5 Scoring rules of the model contract

The written specification defines how nested count thresholds, such as 2 or more and 3 or more symptom types, collapse to one finding, and how the score maps to a probability, a risk band, and a multiple of the average. The scorers do not score a patient when the birth date is missing, when the patient is younger than 18 years on day 0 (the index for Long COVID and the scoring date for the other models), when a code for the target condition is recorded by the scoring date or is undated, or, for Long COVID, when fewer than 90 or more than 455 days have passed since the index. Only records dated on or before the scoring date count. The scorers ignore refuted conditions and entered-in-error records and accept partial FHIR dates, reading a partial record date as the earliest compatible day and a partial birth date as July 1 or the 15th of the month. A failed, forbidden, or truncated FHIR search yields an explicit could-not-score result instead of an absent finding.

[7] Frederick Wolfe, Daniel J. Clauw, Mary-Ann Fitzcharles, Don L. Goldenberg, Winfried Häuser, Robert L. Katz, Philip J. Mease, Anthony S. Russell, Irwin Jon Russell, and Brian Walitt.

2016 revisions to the 2010/2011 fibromyalgia diagnostic criteria. *Seminars in Arthritis and Rheumatism*, 46(3):319–329, 2016. doi: 10.1016/j.semarthrit.2016.08.012.
