## Supplementary material for "Design and Technical Evaluation of a Standards-Based System That Delivers Enclave-Derived Case-Finding Models to Patients and Clinicians": STARE-HI Checklist

### Supplementary File 1. STARE-HI reporting checklist

Design and Technical Evaluation of a Standards-Based System That Delivers Enclave-Derived Case-Finding Models to Patients and Clinicians

Items of the Statement on Reporting of Evaluation Studies in Health Informatics (STARE-HI; Talmon et al., Int J Med Inform 2009;78(1):1-9), with item names as in the explanation and elaboration paper (Brender et al., Appl Clin Inform 2013;4(3):331-358). Section numbers refer to the manuscript. The evaluation is a laboratory evaluation of software on synthetic records, so items about participants and organizational settings are answered for that design.

| Item | Topic | Where reported |
|---|---|---|
| 1 | Title | Title page |
| 2 | Abstract | Structured abstract (Background, Objectives, Methods, Results, Conclusions) |
| 3 | Keywords | Keywords line below the abstract |
| 4 | Introduction | |
| 4.1 | Scientific background | Section 1, paragraph 1 (undercoded conditions, companion analyses) |
| 4.2 | Rationale for the study | Section 1, paragraph 2 (enclave export restrictions, the gap between published and deployed models, available standards) |
| 4.3 | Objectives of the study | Section 2 (three design aims and the technical evaluation) |
| 5 | Study context | |
| 5.1 | Organizational setting | Sections 3.1 and 3.6: a prototype tested on synthetic records only, with no deployment site; Section 5.1 (first limitation) |
| 5.2 | System details and system in use | Sections 3.2 to 3.5 (models, model contract, components, data flow and privacy), Figures 1 to 3, Table 1, Supplementary Sections S1 to S3 |
| 6 | Methods | |
| 6.1 | Study design | Section 3.6: laboratory evaluation with seven checks on synthetic records |
| 6.2 | Theoretical background | Section 3.2 (models fitted from released counts) and the companion analyses it cites; the checks themselves are software verification |
| 6.3 | Participants | No human participants. Section 3.3: 18 synthetic FHIR patients (10 demonstration patients and 8 eligibility boundary cases); Protection of Human and Animal Subjects |
| 6.4 | Study flow | Section 3.6: the counterexample review before the other checks, and the fixes it prompted |
| 6.5 | Outcome measures or evaluation criteria | Section 3.6 and Table 2: tests passed, reference outputs reproduced, expected CDS Hooks cards, SMART sandbox agreement with the demonstration, conformance checks |
| 6.6 | Methods for data acquisition and measurement | Sections 3.3 and 3.6 (test runners, reference implementation, sandbox upload with shifted dates) |
| 6.7 | Methods for data analysis | Section 3.6 (exact comparison of outputs); model validation metrics in Section 3.2 and Table 1 |
| 7 | Results | |

| Item | Topic | Where reported |
|---|---|---|
| 7.1 | Demographic and other study coverage data | Section 4.1 and Table 1 (models and their validation populations); Sections 4.2 and 4.3 (18 patients, 162 reference outputs, 9 models) |
| 7.2 | Unexpected events during the study | Sections 4.3 and 5 (paragraph 5): five defects found by the SMART sandbox comparison and the lesson drawn from them |
| 7.3 | Study findings and outcome data | Sections 4.1 to 4.3, Tables 1 to 3, Figure 2 |
| 7.4 | Unexpected observations | Section 4.3 and Table 3: five failure classes found by the counterexample review, and the fixes |
| 8 | Discussion | |
| 8.1 | Answer to study questions | Section 5, paragraph 1 |
| 8.2 | Strengths and weaknesses of the study | Section 5, paragraphs 2 to 6; Section 5.1 (limitations) |
| 8.3 | Results in relation to other studies | Section 5, paragraph 2 (comparison with deployment frameworks inside one health system) |
| 8.4 | Meaning and generalizability of the study | Section 5, paragraphs 2 to 7; Clinical Relevance Statement |
| 8.5 | Unanswered and new questions | Section 5, paragraphs 4 and 7, Section 5.1, and Section 6 (real-record interoperability, usability, prospective evaluation) |
| 9 | Conclusion | Section 6 |
| 10 | Authors' contribution | Submission form (CRediT roles) |
| 11 | Competing interests | Conflict of Interest |
| 12 | Acknowledgements | Acknowledgments |
| 13 | References | References |
| 14 | Appendices | Supplementary Material (component details and component limitations) and this checklist |
